# Estimating COVID-19 outbreak risk through air travel

**DOI:** 10.1101/2020.04.16.20067496

**Authors:** Y. Daon, R.N. Thompson, U. Obolski

**Affiliations:** School of Public Health, Tel Aviv University, Tel Aviv, Israel; Porter School of the Environment and Earth Sciences, Tel Aviv University, Tel Aviv, Israel; Mathematical Institute, University of Oxford, Oxford, UK; Christ Church, University of Oxford, Oxford, UK

**Keywords:** mathematical model, outbreak resurgence, branching process, policy changes,, Fragile States Index, infectious diseases, imported cases

## Abstract

**Background:** Substantial limitations have been imposed on passenger air travel to reduce transmission of SARS-CoV-2 between regions and countries. However, as case numbers decrease, air travel will gradually resume. We considered a future scenario in which case numbers are low and air travel returns to normal. Under that scenario, there will be a risk of outbreaks in locations worldwide due to imported cases. We estimated the risk of different locations acting as sources of future COVID-19 outbreaks elsewhere.

**Methods:** We use modelled global air travel data and population density estimates from locations worldwide to analyse the risk that 1364 airports are sources of future COVID-19 outbreaks. We use a probabilistic, branching-process based approach that considers the volume of air travelers between airports and the reproduction number at each location, accounting for local population density.

**Results:** Under the scenario we model, we identify airports in East Asia as having the highest risk of acting as sources of future outbreaks. Moreover, we investigate the locations most likely to cause outbreaks due to air travel in regions that are large and potentially vulnerable to outbreaks: India, Brazil and Africa. We find that outbreaks in India and Brazil are most likely to be seeded by individuals travelling from within those regions. We find that this is also true for less vulnerable regions, such as the United States, Europe, and China. However, outbreaks in Africa due to imported cases are instead most likely to be initiated by passengers travelling from outside the continent.

**Conclusions:** Variation in flight volumes and destination population densities create a non-uniform distribution of the risk that different airports pose of acting as the source of an outbreak. Accurate quantification of the spatial distribution of outbreak risk can therefore facilitate optimal allocation of resources for effective targeting of public health interventions.

## Introduction

Coronavirus disease 2019 (COVID-19), caused by the Severe acute respiratory syndrome coronavirus 2 (SARS-CoV-2), has been spreading rapidly since early cases were detected in Wuhan, China at the end of December 2019.^1^ On March 11, the WHO declared the COVID-19 outbreak a pandemic;^2^ i.e. it is now deemed to be on a global scale, with significant autochthonous transmission in different countries. The implications that such a declaration can have are that tackling the COVID-19 pandemic might require international coordination and collaboration.^3^

To date, COVID-19 cases have been observed in over 100 territories, in all inhabited continents.^4^ Such a pervasive spread is partly a consequence of the innate transmissibility of the virus, which is expressed by the relatively high basic reproduction number point estimates ranging from 1.4 to as high as 6.5 (with uncertainty bounds providing an even wider range).^5,6^ However, an additional prominent reason for the especially wide and rapid spread of COVID-19 is likely to be the current high prevalence of international air travel, which has rapidly increased during the last decades. ^7^ Moreover, air travel might have an interaction with the clinical characteristics of COVID-19, increasing the potential of the virus to spread. Although various other infectious diseases have caused epidemics through air travel,^8–10^ none, barring influenza, have had as wide a global spread as COVID-19. Likely factors affecting this significant scale of transmission are the respiratory nature of the disease and a high frequency of presymptomatic (and, probably, to a lesser extent asymptomatic) infectors.^11^ Both factors, which apply to influenza as well,^14^ are hypothesized to help the spread through air travel.^12,13^

Although the spread of COVID-19 through maritime transport has received substantial media coverage due to significant transmission on the *Diamond Princess* and other cruise ships,^14^ most imported cases outside China have likely originated from air travel.^15^ These have spurred the applications of various outbreak control measures regarding air travel, including air travel restrictions, and quarantining individuals returning via air travel from specific countries, or regardless of their departure locations.^16^ Consequently, sharp decreases in flight volumes have been observed,^15^ in contrast with previous projections of a continuous increase in global air travel.^17^ However, such outbreak control measures are non-selective, and are hence costly, inconvenient and substantially hamper general international travel. As such, at least a partial return to high flight volumes is plausible in the future.^18^

A less restrictive possibility to control the COVID-19 spread is to concentrate efforts of detection and isolation of cases to specific locations. Risk assessments of scenarios of COVID-19 spread to new locations, or retrospective analysis or importation to certain locations, have been previously performed.^19–23^ These studies mainly focused on the introduction of COVID-19 into highly vulnerable locations, often with public health infrastructures that may not respond efficiently to contain an outbreak.

On the other hand, locations can also be determined as dangerous by their likelihood to disseminate infected individuals and initiate novel or reoccurring COVID-19 outbreaks in other locations. Investing additional resources in individual screening at high risk source locations might therefore prove efficient in preventing spillover from countries which do not completely cancel international flights. In addition to being relevant currently, such a strategy might also be important when SARS-CoV-2 is suppressed to low-frequency or even undetectable circulation. Under this scenario, narrower public health interventions would be needed, and increased focus would be pointed at preventing new or recurrent outbreaks.

In this study, we consider a possible future scenario in which case numbers are low. We develop and examine a method for discriminating airports with a high potential of initiating a COVID-19 outbreak elsewhere upon the introduction of an infected passenger. We apply this method to modelled flight data, and examine the risk of a new outbreak initiation globally, as well as in three regions where the effects of a sustained COVID-19 outbreak may be especially devastating: Africa, India and Brazil.

Our results are analysed and found robust to various disease outbreak modelling assumptions as well as to seasonal changes in flight patterns.

## Methods

### Data

We approximated monthly data of airport connectivity by the model presented in Mao et al.^24^ Briefly, this data is obtained by applying a Poisson regression model for the number of passengers on covariates such as city connectivity, purchasing power index,^25^ population and weather. This process has yielded information regarding *n =* 1364 major airports scattered throughout the world. The analyses presented in the text rely on air travel during October. This month was chosen as it approximates the beginning of major seasonal respiratory diseases^26^ and is hypothesized to be the potential time of resurgence of COVID-19.^27^ However, similar results are obtained for different months as well (see supplementary material Figure S1). Continent and country information for each airport were retrieved from.^28^

Human population density data was retrieved from NASA.^29^ For each airport *i =* 1,…, *n* an associated density, ρ*_i_* was calculated. First, the location of maximum population density within a 130km radius of each airport was found in order to determine the most likely destination for travellers arriving at that airport. Then, the average population density within a 30km radius of that location was calculated to estimate the density of the population in that area. This heuristic of density estimation is meant to approximate the density of a major nearby location to which an infected passenger is likely to arrive. For example, our heuristic method found the maximal density location of JFK, EWR and LGA (three major airports serving New York City) to be located in downtown Manhattan. Similarly, the two airports serving Berlin (TXL, SXF) and two airports serving Tokyo (NRT, HND) were correctly assigned the same maximal population density in their corresponding metropolitans.

### Outbreak probability

Briefly, we assume a low prevalence of COVID-19 in each country, and use the number of passengers transferring from and to different airports to calculate the probability of arrival of infected individuals to each destination; in each destination, we scale *R* by the local density; calculate the risk of an outbreak using a branching process-based method; and, finally, combine these estimates to yield the risk each airport poses for a COVID-19 outbreak initiation. More details follow.

We used population density estimates to adjust the reproduction number, the expected number of cases generated by a single infected individual in the population, with the rationale being that individual contact rates scale approximately proportional to population density,^30^ which is consistent with another COVID-19 reproductive number estimate.^31^ The reproduction number for region *i, R_i_*, was calculated as *R_i_ = R_w_* • ρ*_i_* /*ρ_w_*, where *ρ_i_* is the density of region *i, ρ_w_* is the estimated density associated with Wuhan Tianhe International Airport, and *R_w_* is the estimated reproductive number in Wuhan. The value of *R_w_* was set to 3 in all the results in the manuscript, as per recent estimates of the basic reproduction number.^5^ A sensitivity analysis for *R_w_* values of 2 and 4 was conducted to reflect the variance in the estimates and potentially lower values in previously exposed populations, and yielded similar results (see supplementary material Figure S3).

After obtaining the basic reproductive number for each location *i*, we can calculate the probability of no outbreak in that location based on the arrival of a single infected individual, *p_i_*, according to the formula derived from a branching process model^32^ *p_i_ =* 1*/R_i_*. We note that this formula remains valid when the population has previously been exposed to the virus - the basic reproduction number can be simply replaced with the effective reproduction number. Such cases are covered by the sensitivity analysis performed on lower basic reproductive number values. Furthermore, this formula is correct for an exponential infectious period, which is often implicitly assumed in infectious diseases dynamic models.^33^ Nonetheless, we performed a sensitivity analysis on the general case of a gamma distributed infectious period, with the appropriate correction,^32,34^ while varying the shape parameter from the exponential case *k =* 1 to a a high value of *k =* 6 (see supplementary material Figure S2 for details). Results were qualitatively similar.

In addition, we have performed a sensitivity analysis for the existence of superspreaders.^35^ The derived formula for *p_i_* in this case is more complex (see derivation in supplementary material), but our results remained robust to this generalization as well (see supplementary material Figure S4).

To calculate the risk each airport poses to an initiation of a COVID-19 outbreak, we assume a fraction *q* of the population is infected. We assume that *q* is small when our method is used to mitigate risk of outbreak initiation or resurgence, probably in the order of magnitude of 10 ^−5^ − 10 ^−6^, since otherwise control measures including complete flight restriction are likely to be implemented. We denote the number of individuals travelling from airport *k* to airport *i* by *n_k,i_*. Therefore, the probability of **no** outbreak initiation from airport *k* (to any airport it is connected to) is 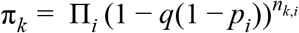 where *i* goes over the range of airports connected to airport *k*. Since *q* is small, this is well approximated by 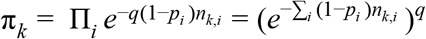. As raising to a positive power *q* preserves order (*a > b* >0 implies *a^q^ > b^q^*), we can conclude the relative order of risk between two airports *k* and *l*, *π_k_* and *π_l_*, does not depend on *q* (as long as *q* is small enough to preserve the approximation above). This independence of *q* enables us to rank the risk of outbreak initiation, 1 *− π_k_*, between different airports regardless of the value of *q*. For ease of presentation, in our results we choose a value of *q =* 10^-6^ yielding low numbers of infected individuals.

Lastly, we turn to highlight three low-middle income regions with high FSI values; Africa, India and Brazil. For each region, we estimated the probability of an outbreak conditioned on infected individuals arriving in an airport in that region. The calculation of the probability of arrival is identical to the one described above, with the exception of restricting flights only to those destined to said regions.

Additional results are provided in the supplementary material, including a table of airport risks corresponding to Figure 1.b (TableS1), a table of airport characteristics (including the associated density, *R* and probability of outbreak following the introduction of one infected individual - all in TableS2), and a full table of the risks by different airports (TableS3). Code and detailed instructions for running the model are available at https://github.com/yairdaon/infections. Code was run using python 3.7.5.

**Figure 1:**
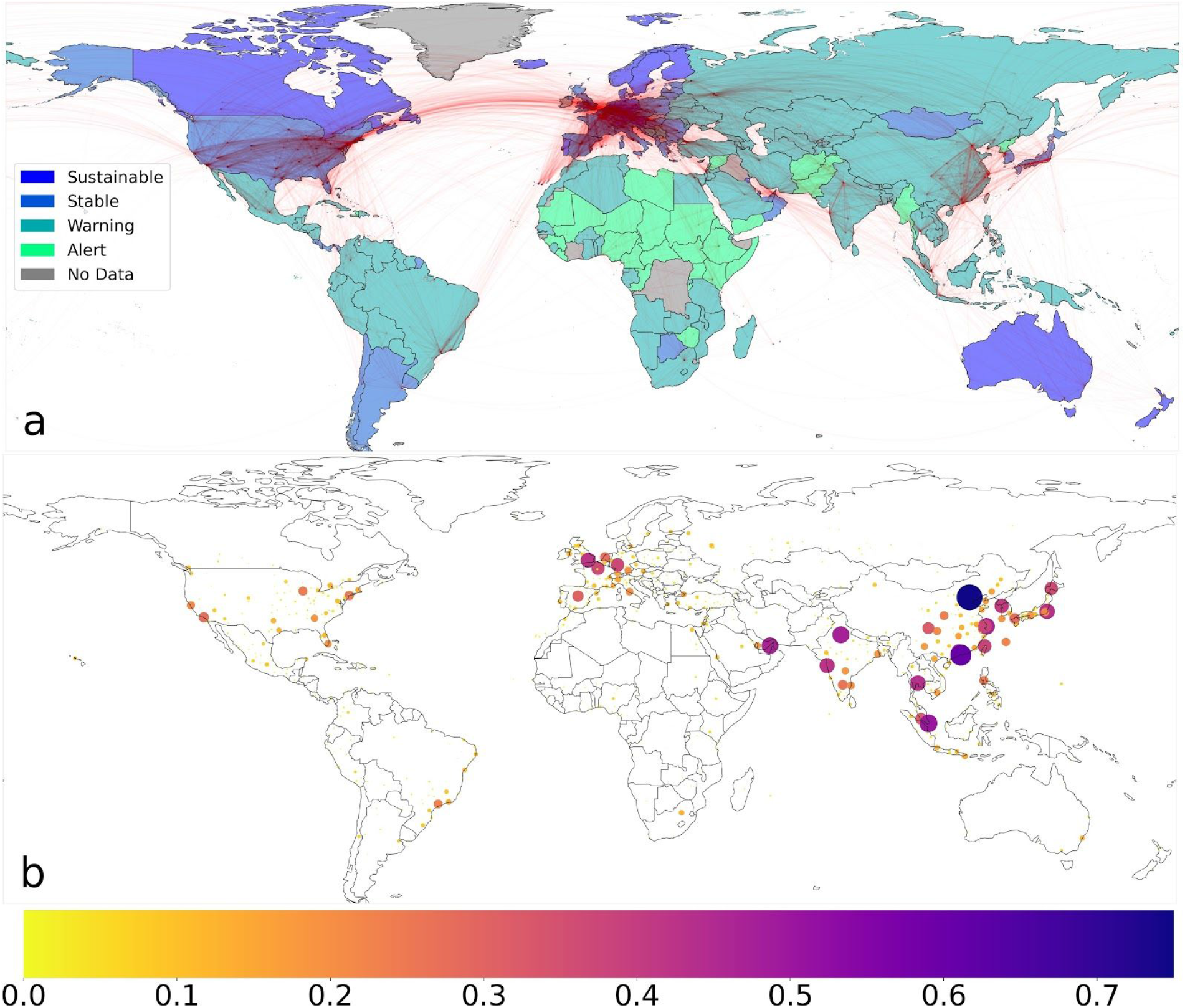
Global airport connectivity and risk of outbreak initiation. a) A global map, overlaid with connections between airports (red) and colored by the Fragile States Index categories^36^. Each connection represents the amount of air travel between locations, to which the curve opacity is proportional. b) The probability of passengers from different airports to initiate a COVID-19 outbreak somewhere around the world, represented by color and circle area. Note that for visualization purposes, the color scale is truncated to [0,0.75].

## Results

We heuristically examine our method by identifying which places have the highest risk of transmission from Wuhan Tianhe International (WUH) Airport. Reassuringly, focal locations to the COVID-19 outbreak appeared among the top-5 airports most likely to have an outbreak originating in WUH, including Beijing, Shanghai and Shenzhen.

We start our analysis by examining the global risk of outbreak by airport. That is, every airport is assigned a risk of initiating an outbreak in any location, according to the airport’s flight patterns and the destination’s risk of outbreak. First, we observe the strong connectivity clusters focused in Europe, east Asia, and the United States (Figure 1.a). We further note that risk is high in east Asia with all but two of the top ten highest-risk airports originating in that region (the exceptions being London Heathrow and Dubai airport; Figure 1). Calculating the average risk of the ten highest-risk airports in each continent yields the following risk order: Asia (0.50), Europe (0.26), North America (0.19), South America (0.11), Africa (0.06), and Oceania (0.03). The estimates of risks in our results greatly vary between airports. For example, the risk estimated for Beijing Capital airport - the airport with the highest risk to initiate an outbreak - is approximately 0.74. On the other hand, Paris Charles De Gaulle airport, ranked 15th, has a risk estimate which is lower by a factor of two.

The locations most vulnerable to be severely affected by a COVID-19 outbreak can be estimated by the Fragile States Index (FSI),^36^ and are mostly low-middle income countries^22^ (Figure 1.a). Hence, we turned our focus to examine the risk of initiating an outbreak in three locations of high FSI values and large populations: Africa, India and Brazil.

Africa is relatively disconnected from the rest of the world. Its strongest connections to locations outside the continent include connection to central Europe, with a relatively strong connectivity to France (mainly through Morocco), England, and the Arab peninsula (Figure 2.a). The risk posed by different airports to initiate a COVID-19 outbreak in Africa is highest at Johannesburg, Dubai, Paris, London, Abuja (Nigeria), Jedda (Saudi Arabia), Frankfurt, Amsterdam, Nairobi and Cairo (Figure 2.b). Contrary to Africa, in India we do observe the largest risk to initiate an outbreak arising from domestic flights, originating within India itself. Of the ten highest-risk airports with respect to India, eight are within the borders of India (the exceptions being airports in Dubai and Singapore). The same trend continues in the following twenty highest-risk airports - fifteen of the top twenty are in India. In Brazil, a similar trend is observed - the 12 highest-risk airports are within Brazil (Figure 4).

**Figure 2:**
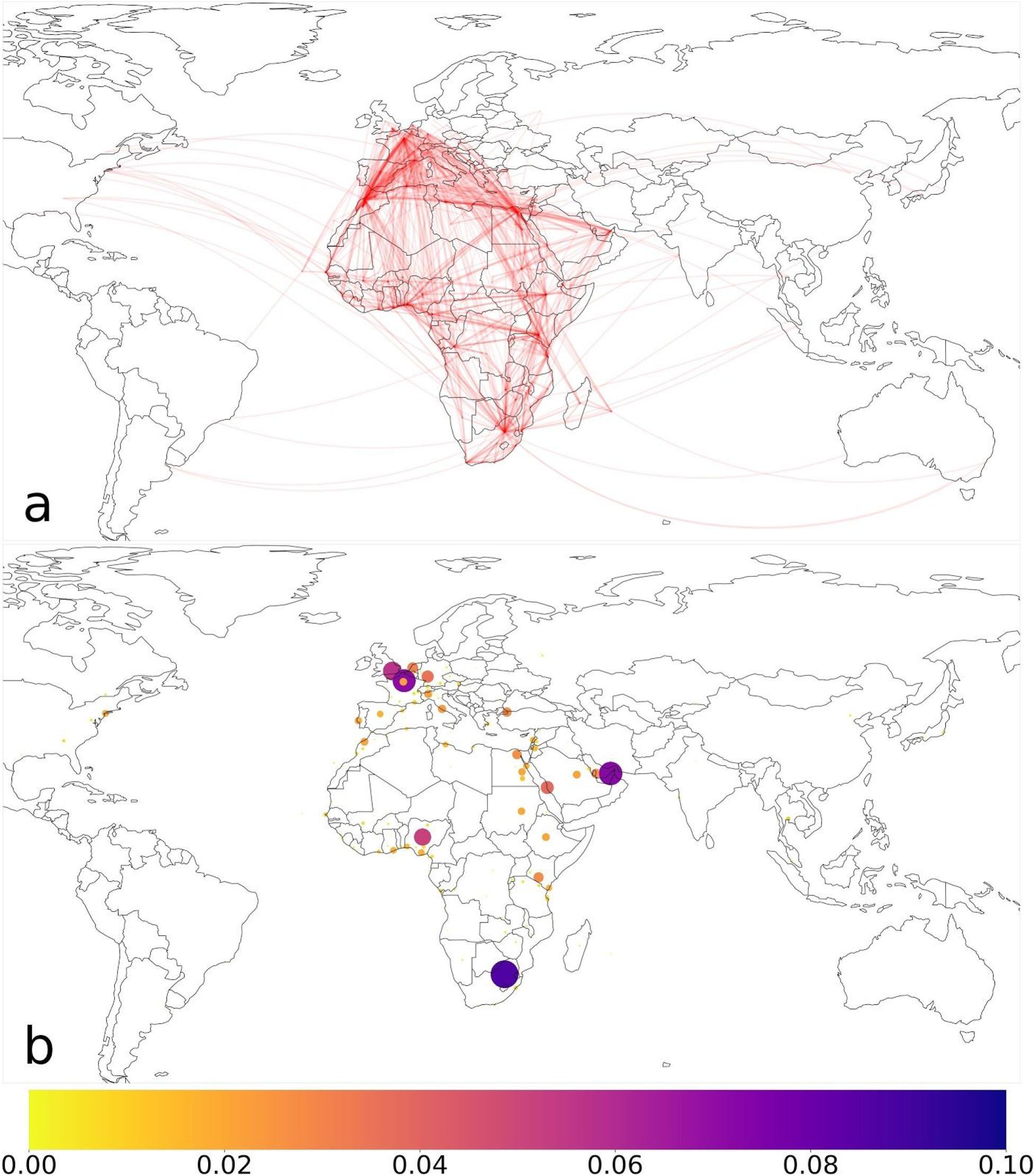
Airport connectivity and risk of outbreak initiation in Africa. a) A global map, overlaid with connections between global airports to an airport in Africa (red). Each connection represents the amount of air travel between locations to an airport in Africa, to which the curve opacity is proportional. b) The probability of passengers from different airports to initiate a COVID-19 outbreak in Africa, represented by color and circle area. Note that for visualization purposes, the color scale is truncated to [0,0.1].

**Figure 3:**
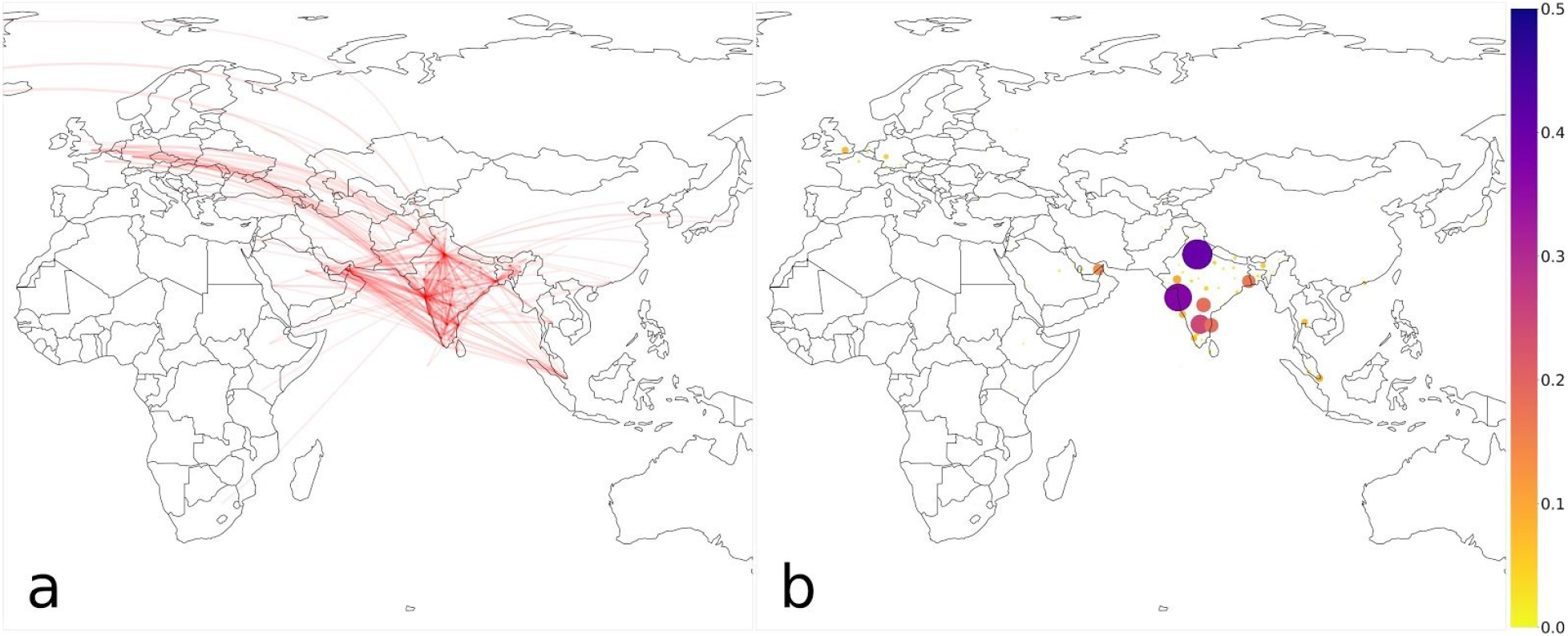
Airport connectivity and risk of outbreak initiation in India. a) A global map, overlaid with connections between airports worldwide to airports in India (red). Each connection represents the amount of air travel between global airports to an airport in India, to which the curve opacity is proportional. b) The probability of passengers from different airports to initiate a COVID-19 outbreak in India, represented by color and circle area. Note that for visualization purposes, the color scale is truncated to [0,0.5].

**Figure 4:**
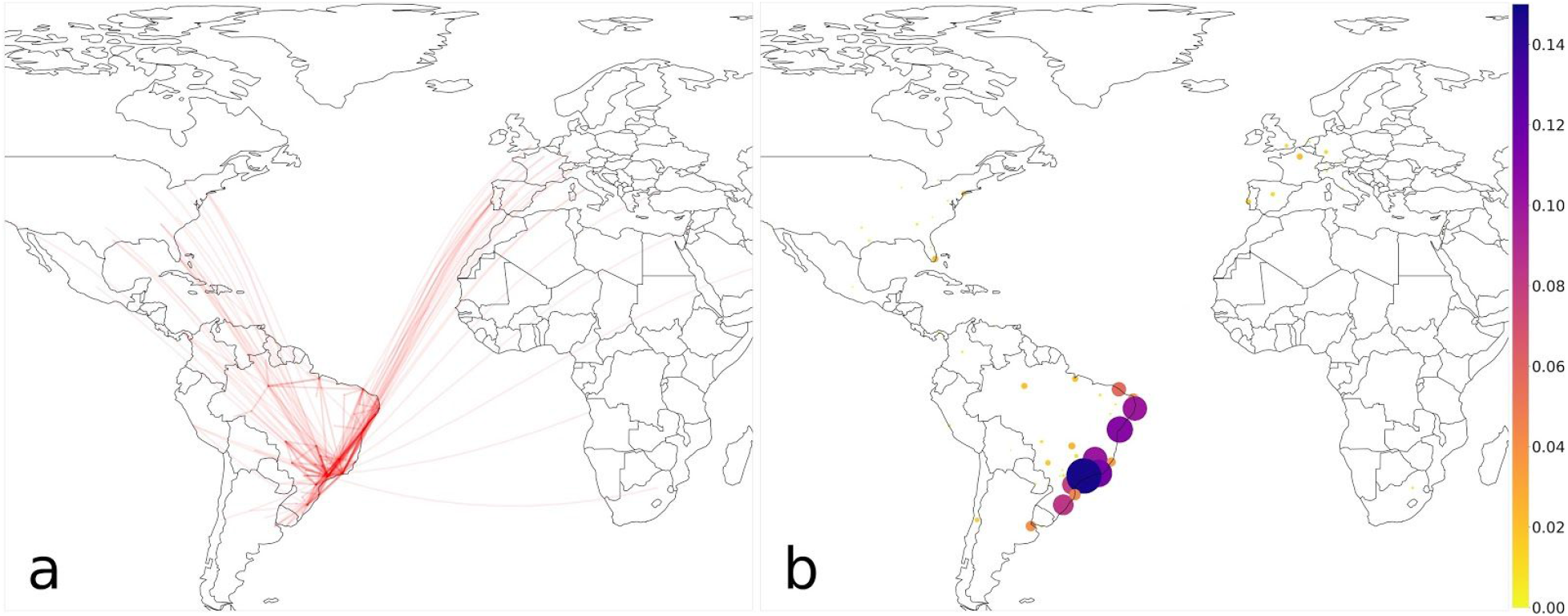
Airport connectivity and risk of outbreak initiation in Brazil. a) A global map, overlaid with connections between airports worldwide to airports in Brazil (red). Each connection represents the amount of air travel between global airports to an airport in Brazil, to which the curve opacity is proportional. b) The probability of passengers from different airports to initiate a COVID-19 outbreak in Brazil, represented by color and circle area. Note that for visualization purposes, the color scale is truncated to [0,0.15].

The trend of highest risk posed by airports within a region persists when considering other large regions - China, the United States, Russia and Europe (supplementary figures S6, S7, S8 and S9, respectively): The risk of an outbreak in Europe arises almost exclusively from airports within Europe, with only three of the thirty highest risk airports outside of Europe. The United States displayed a similar trend; all of the thirty highest-risk airports but five were found within the United States, the exceptions being London, Toronto, Paris, Frankfurt and Tokyo. As far as China is concerned, risk was due mostly to airports in East Asia, with all top forty airports that pose the highest risk to China in said region. Of these, only five are located outside of China (Hong Kong, Seoul, Tokyo, Singapore, Taipei and Bangkok). Russia presents a similar pattern, but to a lesser extent. Four of the ten airports ranked of highest risk to Russia are outside its borders. Three of the rest are in western Europe while one is in Ukraine. We further investigated the seasonal changes in the connectivity between airports. These did not yield substantial differences (see supplementary material Figure S1).

## Discussion

Our study highlights the importance of quantifying the risk of COVID-19 outbreaks through the introduction of cases from air travel. We show that (a) East Asia is the area most prone to initiating an outbreak, globally; (b) Large regions (India, Brazil, the United States, Europe, and China) are at a higher risk for outbreaks from infected passengers arriving from within rather than outside these regions. Although, Russia is somewhat of an exception to this rule; and (c) Africa is a marked exception to this phenomenon, as outbreaks in there are more likely to be initiated by infected passengers from West Europe rather than from within Africa.

Our work complements the results of a recent study that showed that the spread of COVID-19 in Europe closely followed air travel patterns and that the severe travel restrictions implemented there resulted in substantial decreases in the disease’s spread.^37^ That study has demonstrated that such restrictions often came into effect too late after the reporting of COVID-19 cases. In accordance, our study emphasizes that implementation of strict epidemiological control measures to prevent the spread of COVID-19 is likely not to be feasible or timely. We suggest that an additional layer of control measures, such as thorough screening of infected hosts, is important, and that particular attention should be paid to airports that are estimated as likely to be a source of COVID-19 outbreaks elsewhere. Not only does such a strategy prevent a potential outbreak at the destination, but contacts between infectious and susceptible hosts on flights are less likely to occur compared to a strategy based on screening individuals on arrival in a new location.

Unfortunately, non-laboratory based methods of COVID-19 infection detection that are both rapid and accurate are still lacking, and airport screening currently relies on manifestation of symptoms (e.g. body temperature). Hence, individuals with asymptomatic or paucisymptomatic infections may still be missed despite high coverage rates of screening at airports, until newer methods of screening are developed. Current estimates for the frequency of such individuals and their infectivity are still uncertain, ^38,39^ but high frequency of such individuals may regardless force the introduction of new control measures.^35,40^

The transmission of various viral pathogens is affected by seasonal changes in temperature and humidity.^41^ Unfortunately, such effects have yet to be accurately estimated for SARS-COV-2, although some initial investigations in this direction have been made recently.^42–44^ Hence, our study did not consider the potential effects that weather can have on the estimated risk. However, if such changes will be estimated in the future, they could be easily incorporated in our framework, by scaling the different *R* values according to the estimated effects of weather conditions in each location, and recalculating the risk. This could potentially change some of the risk estimates we present, especially of regions mostly connected to either the Northern or Southern hemisphere. In such regions, the differences in weather are expected to be the greatest in different seasons, and these are indeed often expressed in different seasonality patterns of infectious diseases.^45^

Application of the risk estimation we perform, and other assessments relying on air travel data, can be utilized in real-time only if flight travel data are publicly available and easily accessible. Unfortunately, we were not able to obtain such data, and our model relied on previously modelled estimates (see Methods).

We also note that our data do not distinguish between connecting and direct flights, which could affect the estimates of outbreak probabilities around airports that are mostly used as transfer points rather than final destinations of passengers. Some of this effect could perhaps be mitigated by the transferring individuals interacting and infecting workers at those airports, but unfortunately this is outside the scope of our work and data to refine this point are unavailable to us.

Moreover, we did not apply our estimates to other modes of transportation. Rail travel, in particular, could be another relevant route of COVID-19 case importation. India, for example, has more than 10-fold annual train passengers than air passengers,^7,46^ although this figure can vary substantially across countries.^47,48^ We expect rail travel to mostly increase the chances of domestic case importation, due to its more local routes relative to air travel. Potentially, our method could be similarly applied to rail travel in the future, given rail travel data.

Therefore, we emphasize the need in recent, detailed transportation data, and its importance to facilitate research and improved decision making for international COVID-19 outbreak control. Such data are crucial for real time management of crises of a scale we are currently observing.

To conclude, our method provides an estimate of the risk to initiate a COVID-19 outbreak due to travel from different airports. We note that, while connectivity between different locations is important, it is only one component determining the risk of an outbreak. Other important factors include the reproduction number and characteristics of the destination such as population density. Our risk measure combines connectivity weighted by reproduction number estimates in a non-linear fashion, giving rise to a realistic risk measure for outbreaks. Our results demonstrate the heterogeneity in the risk of outbreak initiation on both global, as well as more local scales. As numbers of cases of COVID-19 worldwide decrease, the current shutdown of air travel will be reduced due to the large economic costs. At that time, we suggest that control and screening measures are implemented carefully, with particular attention paid to locations that might be responsible for outbreak initiation.

## Data Availability

All data used are available in the supplementary material and in a github repository.

https://github.com/yairdaon/infections

## Contributions

YD and UO designed the study, analysed the results and wrote the first draft of the manuscript; YD has implemented the analysis; YD, RNT and UO wrote the final manuscript.

